# Reproducibility of Survey Measurements of Sexual Orientation in the USA

**DOI:** 10.1101/2022.04.12.22273781

**Authors:** Charles F. Turner, Brett A. Turner, James R. Chromy

**Affiliations:** City University of New York, Queens College and the Graduate Center, Flushing, New York, United States of America; City University of New York, Queens College, Flushing, New York, United States of America, Current address: Harvard T.H. Chan School of Public Health, Boston, Massachusetts, United States of America; RTI International (Emeritus), Research Triangle Park, North Carolina, United States of America

**Keywords:** survey design, bisexuality, sexual orientation, sexual identity, sexual minorities

## Abstract

Survey measurements of sexual orientation have become increasingly common in national population surveys although validation of these measurements is rare and inherently problematic. We instead assess the *reproducibility* of parallel measurements from two independent samples of the USA population made in the 2008-2018 General Social Surveys and the adult probability subsets of the 2013-2018 National Health Interview Survey (Ns = 12,098 and 190,113).

Restricting analysis to the categories gay/lesbian, bisexual, and straight, we obtain similar estimates of the proportion of the U.S. population who consider themselves gay/lesbian (NHIS: 1.59% vs. GSS: 1.93%, p = 0.059) but not bisexual (NHIS: 1.03% vs. GSS: 2.90%, p < 0.001). Fitting multinomial logistic regression models controlling for year, gender, birth cohort, education, and race, we find that compared to the NHIS, the GSS had 1.248 (p=0.022) times higher relative odds of eliciting a response of Gay-Lesbian (vs. Straight) and 2.980 (p<0.001) times higher relative odds of eliciting a response of Bisexual (vs. Straight). Expanding the model by adding 3-way interaction terms for orientation-by-predictor-by-survey, we find that we cannot reject the null hypothesis that *trends* over time and across subpopulations in reporting of sexual orientation were statistically equivalent for the two survey programs.

## 1. Introduction

Since the 1990s, survey measurements of sexual orientation have become increasingly common in population surveys of sexual behavior in the USA, Europe, and elsewhere [1-4]. Assessments of the accuracy of such measurements are infrequent, however, and can be problematic.

There is a large literature documenting the ways in which seemingly trivial variations in survey protocols can induce substantial discrepancies in the measurements obtained. These include seemingly minor variations in question wording, context, mode of administration, etc. [5-7]. This problem has been long known to affect so-called “objective” measurements (e.g., personal bankruptcy [8]) in which the notion of a measurement’s “true value” can be logically sustained. In those instances it is theoretically possible to attempt validation by comparing survey reports to a presumed veridical record (e.g., comparing survey reports of personal bankruptcy to court records). This is not to say, of course, that such comparisons can be easily (or ever) accomplished for every objective phenomenon; consider for example the difficulty of validating reported condom use at last sex. While partner reports might seem to provide a reasonable strategy, there are inherent limitations to this approach, e.g., in the case of anonymous hook-ups.

This problem becomes even more difficult for subjective measurements where there is simply no veridical record that can be used to validate survey reports of: how happy someone is, their approval of the President, or their sexual orientation [9]. A preliminary question, however, can be fruitfully attacked without taking on the epistemological challenges posed by “validation”. That question is the *reproducibility* of the survey measurements of the subjective phenomena that is at issue. For us it is the reproducibility of survey measurements of sexual orientation.

In the present instance we have the benefit of two independent series of surveys that measured the reported sexual orientation of the U.S. adult population. These measurements come from the 2008 to 2018 waves of the biennial General Social Survey [10] and the 2013 to 2018 waves of the annual National Health Interview Survey [11,12]. These data sources have two important advantages for assessing the reproducibility of survey measurements of sexual orientation. First, they are well-regarded surveys of large probability samples of the U.S. population (Ns across available years =12,098 GSS, and 190,113 for the probability adult segment of the NHIS samples; it should be noted that while the GSS conducted a total of 13,794 interviews in 2008 through 2018, only 12,098 respondents were included in survey segments that included the Sexual Orientation question.) Second, these survey programs were conducted by well-trained staffs of survey interviewers and other survey professionals.

*The NHIS program* is the USA’s premier health interview survey. Begun in 1957, the NHIS is designed “to secure accurate and current statistical information on the amount, distribution, and effects of illness and disability in the United States and the services rendered for or because of such conditions” (https://www.cdc.gov/nchs/nhis/about_nhis.htm, accessed January 13, 2020). This survey program annually draws large representative samples of the USA population and uses U.S. Census Bureau interviewers to conduct interviews. For example, in the 2018 survey, 29,839 households were included in the NHIS sample [11, p. 20]. One probabilistically selected adult from each sampled household was selected for inclusion in the adult probability component of the NHIS. In 2018 that yielded 25,417 completed interviews. This component of the 1997 through 2018 NHIS is the focus of all of our NHIS analyses (unweighted annual Ns range from 21,781 to 36,697).

*The GSS Program* is a major representative sample survey of adults residing in households in the USA. It has been conducted since 1972 as part of the National Science Foundation’s National Data Program for the Social Sciences [10]. The GSS is conducted using in-person interviews conducted by professional interviewers employed and supervised by the National Opinion Research Center at the University of Chicago. GSS surveys were conducted annually between 1972 and 1994 and biennially in subsequent years. These surveys have included “a standard core of demographic, behavioral, and attitudinal questions, plus topics of special interest. Among the topics covered are civil liberties, crime and violence, intergroup tolerance, morality, national spending priorities, psychological well-being, social mobility, and stress and traumatic events” [13].

Beginning in 1988, the GSS has included questions on sexual behaviors; in 2008 a question on sexual orientation was added. The GSS sexual orientation data from 2008 through 2018 provide the basis for our comparative analyses. The 2008-2018 GSS drew probability samples of US households and interviewed one probabilistically selected adult from each household. Biennial GSS samples included between 1,974 and 2,867 adults in 2008 through 2018. (In the 2016 and 2018 GSS the sexual orientation question was only asked of a randomly selected two-thirds of the survey sample.)

## 2. Materials and Methods

### 2.1 Questioning

While the sexual orientation question was quite similar in the two survey programs, there was a minor difference in the question stem and a more substantial difference in the response categories that were offered (see Table 1). There were also substantial differences in the introductory texts and questions preceding the sexual orientation measurement and in the methods of question administration.

**Table 1.** Questions Measuring Sexual Orientation in the 2008-2018 General Social Surveys and National Health Interview Surveys.

| Component | General Social Survey (a,b) | National Health Interview Survey |
| --- | --- | --- |
| Introductory Text | "There is a great deal of concern today about the AIDS epidemic and how to deal with it. Because of the grave nature of this problem, we are going to ask you some personal questions and we need your frank and honest responses. Your answers are confidential. | "You are about to enter the Sexual Identity and Lifestyle questions section. This section includes questions on computer use, the respondent's neighborhood, sexual identity, financial worries, mental health, and HIV testing." |
| Question Wording | Which of the following best describes you? | Which of the following best represents how you think of yourself? |
| Response Categories | Gay, Lesbian or Homosexual<br>Bisexual<br>Heterosexual or straight<br>(Computer provided dedicated keys to indicate "don't know" and "refused".) | 1. Gay (*if male*); Lesbian or Gay (*if female*);<br>2. Straight, that is, not gay (*if male*);<br>Straight, that is not lesbian or gay (*if female*)<br>3. Bisexual<br>4. Something else<br>5. I don't know the answer |
| Mode of Administration | Computer-Assisted Self Interview (Video-CASI) (b) | Interviewed-administered using CAPI *with Flashcards in Male or Female versions.* |

(a) Source: [13].
(b) GSS dataset in 2008 (but not other years) includes variable SCIMODE which is described as "Is the SAQ self administered or interviewer administered". Responses in 2008 were: Assigned and done as interviewed = 1,237; Assigned and done as SAQ = 140; Assigned as SAQ done in interview due to R's request/inability to do as SAQ = 89; Assigned as SAQ done in interview on phone = 39; Inapplicable = 518.

Using video-CASI, the GSS administered the sexual orientation question and a range of questions on sexual behaviors including a variety of questions on number, gender, and types (friend, spouse, date, sex worker, etc.) of sexual partners in lifetime and during recent years (1, 5, etc.), frequency of sex, condom use, injection drug use, cocaine use, etc. [14]. In the video-CASI procedure, interviewers do not ask the survey questions, but rather the questions are shown on the screen of a laptop computer and respondents reply using the laptop computer’s mouse or keyboard. Thus the interviewers are unaware of the questions being asked and the responses the interviewee gives. Dedicated keys on the laptop’s keyboard (as well as categories shown on the laptop screen) allow respondents to answer “Don’t Know” or to refuse to respond to the question)

A number of randomized survey experiments ([15-17] report substantial differences in the reporting of sensitive behaviors when CASI survey technologies are used in place of human interviewing. However, a carefully conducted, randomized experiment by NCHS [18] did not find substantial differences in responses to the NHIS sexual orientation question using audio-CASI versus the NHIS “showcard” protocol described here.

### 2.2 Response Categories

The GSS sexual orientation question asks which of the offered response options “best describes you” while the NHIS sexual orientation question similarly asks which option “best represents how you think of yourself”. The response categories, however, diverged more substantially. The GSS offered the response categories: Gay, Lesbian or Homosexual; Bisexual; and Heterosexual or straight. The NHIS, however, also offered the explicit response categories: “4. Something else” and “5. I do not know the answer” while the GSS does not explicitly offer similar options.

36 of the 12,098 respondents in the 2008-2018 GSS were coded as responding “Don’t Know” (weighted 0.31%, se = 0.05%) on the sexual orientation question. In contrast, 1,259 of 190,113 of NHIS respondents chose the *offered* option of “5 --I do not know the answer” (weighted 0.67%, se = 0.03%). A further difference is found for the “something else” response category. The GSS did not offer this option forcing respondents to choose among the three substantive response categories or to provide no response. In the NHIS, 636 of 190,113 respondents chose the offered answer category: “4 --. Something else” (weighted 0.32%, se = 0.02%).

### 2.3 Differences in the Population Surveyed

While both survey programs provide broadly representative samples of the adult population of the USA, one minor divergence should be noted. The GSS sample is limited to the *household* population while the NHIS sample covers the entire civilian non-institutionalized population. So, for example, the NHIS includes a dormitory component in its sampling frame while the GSS does not. As a crude check of the impact of this variation, we compared NHIS estimates of sexual orientation with and without the non-household segment of the NHIS sample. Since only 0.95% of the NHIS adult probability sample (unweighted N = 1,739 of 183,020) were non-household residents, it is not surprising to find (see Appendix Table B) that the estimates are virtually identical.

Since language barriers are a potential reason for non-response in both survey programs, it is important to note that both survey programs conducted interviewing in both English and Spanish in 2008 -2018.

### 2.4 Combining Samples

For our reproducibility analysis, we analyze a combined data set containing data from 6 biannual GSS surveys (2008, 2010, 2012, 2014, 2016, and 2018) and from 6 annual NHIS surveys (2013 to 2018).

The combination of these files is not as straightforward as merely concatenating the two data files and ensuring that variable names and response categories are made equivalent. While that would be sufficient for two simple random samples of the population, it unfortunately is not adequate for these two complex probability samples. In particular, we must ensure consistency in the handling of weights and the variables required for calculation of the effects of the complex sample designs used in each of these survey programs. (Weights include adjustments for sampling factors and in the NCHS post-stratification adjustments for the impact of survey nonresponse. The GSS did not employ such post-stratification adjustments for non-response [19].)

For the analyses reported in this article, a composite GSS-NHIS data file was created. First *new weights* were constructed for each wave of the NHIS (2013, 2014 … 2018) and GSS survey (2008, 2010… 2018). These new weights for survey respondents were computed as the originally assigned weight divided by the mean of all weights for that wave of the survey. As a check on the accuracy of this step, we inspected all permutations of Survey-by-Wave to ensure that the sum of the new case weights equaled the unweighted sample sizes for each of these permutations — as well as for the total survey samples. Strata and PSU numbering were unaltered from the original survey datasets.

### 2.6. Statistical Analyses

We begin our analyses by reporting the number and percentages of GSS and NHIS respondents who were recorded as choosing each substantive response plus those providing non-substantive responses, e.g., refusals to answer. We then attempt direct comparisons of response distributions obtained in the two survey programs using alternative methods for handling discrepant responses in the two survey programs, e.g., the lack of an explicit “Don’t Know” response category in the GSS.

Subsequently, we test the congruence of the results of the two survey programs in monitoring trends in reported sexual orientation over time and birth cohorts and across sociodemographic subsets of the population. All analyses were performed using Stata version 15 [20] and employ Stata’s *svy* procedures to take account of the complex sample designs of these surveys in estimating results for the U.S. adult population.

## 3. Results

### 3.1 Full Response Distributions

The first panel of Table 2 shows results for the full set of response categories for both the GSS and NHIS measurements across all years in which the questions were asked, i.e., annually in 2013-2018 in the NHIS and biennially in 2008-2018 in the GSS. For this initial analysis, we will not synchronize the time periods used in the two surveys.

**Table 2.** Estimated Prevalence of sexual orientations reported by adult population in 2013-18 NHIS and 2008-18 GSS [d].

| SURVEY |  | Gay/<br>Lesbian | Straight | Bisexual | Something<br>Else [a] | Don't<br>Know | Unknown /<br>Not<br>Ascertained<br>[b,c] | Refused | Unweighted<br>Ns |
| --- | --- | --- | --- | --- | --- | --- | --- | --- | --- |
| NHIS | *est.* | 1.58% | 93.79% | 0.93% | 0.32% | 0.67% | 2.19% | 0.51% | 190,113 |
|  | *se* | 0.04% | 0.09% | 0.03% | 0.02% | 0.03% | 0.53% | 0.02% |  |
| GSS | *est.* | 1.50% | 85.32% | 2.00% | na | 0.31% | 9.510% | 1.37% | 12,098 |
|  | *se* | 0.11% | 0.48% | 0.14% | na | 0.05% | 0.430% | 0.13% |  |
| Estimates excluding "Not Ascertained" Responses |  |  |  |  |  |  |  |  |  |
| NHIS | *est.* | 1.62% | 95.90% | 0.95% | 0.33% | 0.69% | na | 0.53% | 186,048 |
|  | *se* | 0.04% | 0.07% | 0.03% | 0.02% | 0.03% | na | 0.02% |  |
| GSS | *est.* | 1.66% | 94.28% | 2.21% | na | 0.34% | na | 1.51% | 10,894 |
|  | *se* | 0.12% | 0.26% | 0.16% | na | 0.06% | na | 0.15% |  |

(a) The "something else" category was only offered explicitly in the NHIS surveys.
[b] The NHIS surveyors report that "For partially completed interviews (e.g., Sample Adult interviews where the respondent discontinued the interviews any time after completing the first three sections), the responses will appear as 8's (not ascertained) for the remaining variables in the file ... A code of 8 is also used to indicate "not ascertained" responses when the field was blank or contained an impossible code. [11, p. 19]" The authors also note that "In the 2018 data file, for example, 3.0% of completed sample adult cases were partially completed interviews that were deemed sufficient for analysis (that is, the sample adult discontinued the interview but did complete at least the first three sections) [11, p. 81)".
[c] The 2018 GSS Codebook notes that "People who did not do this [sex] supplement are coded as IAP." In 2008-2014 the sexual orientation question was asked in all three of randomly-assigned "ballots" of the GSS, and 863 of 8,579 respondents (10.1% unweighted) were coded as IAP in those years. It should be noted that in 2016 and 2018, the sexual orientation question was asked in only two of the three GSS "ballots," and thus all respondents in the excluded ballot were coded as IAP. (These respondents are not included in the tabulation above.)
[d] Estimates shown in Table 2 were calculated from the public use datasets released by the IPUMS Health Surveys website[12] and the NORC General Social Survey website [10].

Not surprisingly, we find that there is notable disagreement in the results obtained by these two measurement programs. A primary source of this disagreement arises from the much higher population percentage that was not asked the sexual orientation question because either:

> In the NHIS, they discontinued the survey before reaching the sexual orientation question (2.19%), or
>
> In the GSS, they discontinued the survey before reaching the self-administered sexual behaviors section of the GSS survey or did not complete this section due to refusal or some other reason and thus were coded as not ascertained (IAP, 9.51%). (The GSS Director has noted that “[the GSS dataset] does not have a variable that delineates refusal to do the SAQ [self-administered questionnaire], although that is what most of the … IAP probably are (Tom Smith, personal communication, March 4, 2020)”.

The relatively large percentage of GSS respondents refusing the sexual behavior section of the survey may reflect the wide range of sensitive information that was requested by this GSS section, e.g., number, gender, and types of sexual partners, frequency of sex, etc. This variation in measurement mechanics perturbs all other results. This is troublesome given our focus on a relatively rare characteristic of the population, i.e., the prevalence of non-heterosexual orientations.

As a rough correction for this measurement variation, the lower panel of Table 2 shows estimated prevalence based only on respondents who were asked the sexual orientation question. Both survey programs estimate that 94% to 96% of the U.S. population describes themselves as heterosexual or straight while approximately 1.6% to 1.7% of the population describes itself as gay or lesbian. An additional 0.5% to 1.5% of the population is classified as refusing to respond to the sexual orientation question. There is, however, less agreement on estimates of the population classifying themselves as bisexual. Although the percentages are low, the GSS estimates more than double the prevalence of self-reported bisexuality in the U.S. population compared to the NHIS survey program (GSS: 2.21%, se = 0.16%; NHIS: 0.95%, se = 0.03%; p<0.001)

Some discrepancies in these estimates might be expected given the variation in the protocols used by the two survey programs. We note, for example, that the NHIS program included an explicit category (“something else”) for respondents who did not feel comfortable using the three sexual orientations: straight, gay-lesbian, or bisexual. In addition, the mode of survey administration varied between the two survey programs.

In addition, we note differences in question wording and administration. NHIS interviewers asked the question, “Which of the following best represents how you think of yourself?” with respondents choosing their response category from a flashcard. In contrast, the GSS program used video-CASI to administer the sexual orientation and other sensitive questions. (A small number of GSS sexual orientation measurements were made by telephone. In 2008, 39 of these GSS measurements were made in interviewer-administered telephone interviews.) In the video-CASI mode of administration, the survey interviewer does not ask the question nor hear the respondent’s choice of a response. This variation in survey mode may have affected the measurements by, among other things, affecting respondents’ willingness to provide an accurate report of their sexual orientation.

It is also possible that variations in survey administration may have affected the salience of non-substantive response options. In the GSS video-CASI administration, respondents were told to press one of two keys on their laptop if they either did not know the answer to the question or did not wish to provide an answer. In contrast, NHIS respondents were offered an explicit option on the flashcard of “I do not know the answer” but no explicit refusal option. These and other variations in survey protocol might account for the incomplete equivalence in the point estimates obtained by the two survey programs shown in Table 2.

A common analytic response to non-equivalence in the response categories offered by two measurement programs is to focus analysis on the common substantive responses. In the present case an analyst might keep the three substantive response categories (straight, gay-lesbian, and bisexual) and drop all cases in which respondents gave a non-substantive response, i.e., don’t know, “something else”, or refused to respond. As shown in Table 2 (lower panel), estimates for these non-substantive response categories total 1.55% for the NHIS and 1.85% for the GSS. While these are fairly trivial percentages, it should be borne in mind that the estimated size of the (self-described) gay/lesbian and bisexual subpopulations are only 3 to 4%.

### 3.2 Simple Analyses Restricted to Three Substantive Response Categories

As shown in Table 3, restricting analysis to the three substantive categories of gay/lesbian, straight, and bisexual, produces quite similar estimates of the proportion of the U.S. population who consider themselves gay or lesbian. The NHIS yields an estimate of 1.59% (se=0.06%) which is only modestly different from the estimate produced by the GSS (1.93%, se=0.17%), and that difference is of borderline statistical significance (p = 0.059). Approximately similar results are found when males and females are considered separately. For males, the NHIS estimates that 1.81% (se=0.09%) identify themselves as gay while the GSS estimates this percentage as 2.13% (se=0.28%; p = 0.277). For females, similar results are obtained with the NHIS estimating that 1.39% (se=0.08%) of females identify as lesbian and the GSS estimating this percentage as 1.77% (se=0.22%, p=0.105).

**Table 3.** Estimated prevalence of self-described sexual orientations of U.S. adults in 2014, 2016, and 2018 NHIS and GSS --- restricting response to three substantive categories of sexual orientation [a].

| SURVEY<br>& Gender |  | Gay/Lesbian | Straight | Bisexual | Unweighted<br>Ns | *P inter-survey*<br>*comparison* |
| --- | --- | --- | --- | --- | --- | --- |
| *Males & Females* |  |  |  |  |  |  |
| NHIS | *est.* | 1.59% | 97.38% | 1.03% | 91,760 | <0.001 |
|  | *se* | 0.06% | 0.08% | 0.05% |  |  |
| GSS | *est.* | 1.93% | 95.17% | 2.90% | 5,421 |  |
|  | *se* | 0.17% | 0.33% | 0.26% |  |  |
| Total | *est.* | 1.62% | 97.22% | 1.16% | 97,181 |  |
|  | *se* | 0.06% | 0.07% | 0.05% |  |  |
| *Males only* |  |  |  |  |  |  |
| NHIS | *est.* | 1.810% | 97.570% | 0.620% | 41,465 | <0.001 |
|  | *se* | 0.09% | 0.11% | 0.06% |  |  |
| GSS | *est.* | 2.13% | 96.35% | 1.52% | 2,464 |  |
|  | *se* | 0.28% | 0.39% | 0.24% |  |  |
| All Males | *est.* | 1.83% | 97.49% | 0.68% | 43,929 |  |
|  | *se* | 0.09% | 0.10% | 0.05% |  |  |
| *Females only* |  |  |  |  |  |  |
| NHIS | *est.* | 1.39% | 97.19% | 1.42% | 50,295 | <0.001 |
|  | *se* | 0.08% | 0.11% | 0.08% |  |  |
| GSS | *est.* | 1.77% | 94.16% | 4.07% | 2,957 |  |
|  | *se* | 0.22% | 0.50% | 0.42% |  |  |
| All Females | *est.* | 1.42% | 96.97% | 1.61% | 53,252 |  |
|  | *se* | 0.07% | 0.11% | 0.08% |  |  |

However, similar equivalence is not found for estimates of the percentage of the U.S. population identifying as bisexual (p < 0.001). Overall, the NHIS survey program estimates that 1.03% (se=0.05%) of the U.S. population identifies as bisexual while the GSS survey program yields an estimate that is more than twice as high (2.90%, se=0.26%). Roughly similar discrepancies are found when men and women are considered separately with the GSS bisexual estimates being two to three times higher than the NHIS estimates; see Table 3).

It should be noted that this non-equivalence is not eliminated by focusing on the contrast between respondents reporting a “straight” sexual orientation versus those reporting a “non-straight” orientation, i.e., gay/lesbian or bisexual. Overall – as might be inferred from Table 3 – the GSS estimates that a higher percentage of the U.S. population identifies with a “non-straight” orientation than does the NHIS (4.83% vs. 2.62%, p < 0.001).

### 3.3 Testing Reproducibility with Controls for Sociodemographic Factors

We refined and expanded the foregoing analyses by fitting multinomial logistic regression models that predict our 3-category outcome variable (gay, straight, bisexual) as a function of the survey making the measurements plus five control variables. These control variables were: the year the survey was conducted, and the respondent’s gender, birth cohort (in decades), education, and race. The latter four predictors have been shown in previous research to be associated with reporting of alternative sexual orientations and/or behaviors (e.g., Turner et al., 2005; England et al., 2016). For these analyses we restricted our sample to the three years (2014, 2016, 2018) in which both the NHIS and GSS surveys collected respondents’ reports of their sexual orientation.

Table 4 presents the results of this analyses. In this analysis the response “straight” is the base outcome. The entries in this table show the estimated relative odds of the responses: (1) gay vs. straight, and (2) bisexual vs. straight, adjusted for the other predictor variables in the model. Our results indicate that compared to the NHIS, the GSS has 1.248 (p=0.022) times higher relative odds of eliciting a response of Gay-Lesbian (vs. Straight) and 2.980 (p<0.001) times higher relative odds of eliciting a response of Bisexual (vs. Straight). Overall, this result indicates that the discrepancies observed in our simpler analyses (Table 3) are not eliminated by the inclusion of the five control variables nor the restriction of our analysis to the three years in which both surveys made measurements.

**Table 4.** Multinomial logistic regression results: (1) predicting relative log odds and (2) relative odds for reporting a gay or bisexual (vs. straight) orientation as a function of the survey making the measurement controlling for birth cohort and selected sociodemographic variables.

| PREDICTOR | RESPONSE<br>Gay - Lesbian |  |  | RESPONSE<br>Bisexual |  |  | *P*<br>Main Effect<br>(b) |
| --- | --- | --- | --- | --- | --- | --- | --- |
|  | Coef.<br>Relative Log<br>Odds (a) | Relative<br>Odds<br>Ratio (a) | P | Coef.<br>Relative<br>Log Odds<br>(a) | Relative<br>Odds Ratio<br>(a) | P |  |
| **SURVEY** |  |  |  |  |  |  |  |
| NHIS | *ref.* | *ref.* |  | *ref.* | *ref.* |  |  |
| GSS | 0.222 | 1.248 | 0.022 | 1.092 | 2.980 | <0.001 | <0.001 |
| **GENDER** |  |  |  |  |  |  |  |
| Male | *ref.* | *ref.* |  | *ref.* | *ref.* |  |  |
| Female | -0.244 | 0.783 | 0.001 | 0.915 | 2.497 | <0.001 | <0.001 |
| **BIRTH COHORT** |  |  |  |  |  |  |  |
| 1930 & earlier | *ref.* |  |  | *ref.* |  |  |  |
| 1940 | 0.495 | 1.641 | 0.020 | 0.160 | 1.173 | 0.630 |  |
| 1950 | 0.931 | 2.537 | < 0.001 | 0.397 | 1.488 | 0.176 |  |
| 1960 | 1.310 | 3.708 | < 0.001 | 0.804 | 2.235 | 0.005 |  |
| 1970 | 1.189 | 3.282 | < 0.001 | 1.171 | 3.226 | < 0.001 |  |
| 1980 | 1.293 | 3.643 | < 0.001 | 1.941 | 6.964 | < 0.001 |  |
| 1990+ (c) | 1.455 | 4.285 | < 0.001 | 2.492 | 12.080 | < 0.001 | <0.001 |
| **EDUCATION** |  |  |  |  |  |  |  |
| <HS | *ref.* | *ref.* |  | *ref.* | *ref.* |  |  |
| HS Graduate | 0.575 | 1.777 | 0.001 | -0.263 | 0.769 | 0.104 |  |
| BA/BS | 0.890 | 2.434 | <0.001 | -0.125 | 0.882 | 0.473 |  |
| Post-BA | 1.217 | 3.378 | <0.001 | -0.108 | 0.898 | 0.575 | <0.001 |
| **RACE** |  |  |  |  |  |  |  |
| White | *ref.* | *ref.* |  | *ref.* | *ref.* |  |  |
| Black | 0.101 | 1.107 | 0.301 | -0.263 | 0.769 | 0.017 |  |
| Other | -0.326 | 0.722 | 0.018 | -0.297 | 0.743 | 0.063 | 0.009 |
| **YEAR OF SURVEY** |  |  |  |  |  |  |  |
| 2014 | *ref.* | *ref.* |  | *ref.* | *ref.* |  |  |
| 2016 | -0.057 | 0.944 | 0.500 | 0.210 | 1.234 | 0.033 |  |
| 2018 | -0.037 | 0.964 | 0.673 | 0.358 | 1.430 | <0.001 | 0.009 |
| **CONS.** | -5.783 |  | <0.001 | -6.544 |  | <0.001 |  |

(a) Estimates derived by multinomial logistic regression accounting for complex design of samples (F[32, 851] = 23.48, $p < 0.001$ ). Coefficients indicate the estimated relative log odds of reporting Gay-Lesbian (or Bisexual) associated with being in the row category (e.g., female) of the predictor variable compared to the base category (e.g., male) when controlling for other factors in the model. The exponentiated value of this coefficient is the corresponding relative odds ratio. For rare attributes, this value is close to the relative risk ratio.
(b) Test of the null hypotheses that coefficients for categories of sexual orientation by predictor are zero.
(c) The 1990+ cohort includes 22 GSS respondents and 210 NHIS respondents born in 2000.

In passing we note that this analysis replicates many aspects of previous findings. Thus we note monotonic increases across birth cohorts in the relative odds that respondents will choose the categories gay-lesbian and bisexual versus straight. These results are quite startling although they are consistent with past research [21-22]. Thus compared to the 1930s, birth cohort, members of the 1990s cohort have 4.3 (p<0.001) times higher relative odds of reporting a gay-lesbian orientation and 12.1 (p<0.001) times higher odds of reporting a bisexual orientation. We similarly find that compared to men, women have 0.78 (p < 0.001) times lower relative odds of reporting a gay-lesbian orientation but a 2.50 times (p = 0.001) higher relative odds of reporting a bisexual orientation.

Controlling for other factors in the model, higher levels of education were associated with increased relative odds of reporting a gay-lesbian (vs. straight) orientation but a similar result is not found for reporting of a bisexual orientation. Thus persons with a college degree and post-college education had 2.43 and 3.38 (ps<0.001) times higher relative odds of reporting a gay-lesbian (vs. straight) orientation’ A parallel result is not found for reporting a bisexual orientation. Surprisingly, this analysis reveals that reported sexual orientations varied significantly across the three years in which the surveys were conducted when other variables are controlled. Most noticeably, relative to respondents in 2014, respondents in 2016 had 1.23 (p=0.033) higher relative odds of reporting a bisexual (vs. straight) orientation and respondents in 2018 had 1.43 (p<0.001) times higher relative odds of reporting a bisexual (vs. straight) orientation. This analysis also reveals a significant association of race with reported sexual orientation. Respondents of “other” races (i.e., not black or white) had 0.72 lower relative odds than whites (p = 0.018) of reporting a gay-lesbian orientation (vs. straight) and borderline lower relative odds (0.74, p=0.063) of reporting a bisexual (vs. straight) orientation. Black respondents also had 0.77 (p=0.017) times lower relative odds than whites of reporting a bisexual orientation although the difference in relative odds of reporting gay/lesbian orientations was not statistically significantly (1.11, p = 0.301).

### 3.4 Reproducibility of Predictor-by-Orientation Associations

While the observed disagreements in point estimates of the sexual orientations reported by the U.S. population are noteworthy and sociodemographic controls do not eliminate that disagreement, it can be argued that this does not detract from one of the most important uses of such data — monitoring *trends over time and variations across subpopulations* in sexual orientations. Indeed, since the measurement protocols used by the two surveys are not fully equivalent, some non-equivalence in these estimates of sexual orientation should be expected. The measurements may nonetheless provide equivalently reliable and important indicators of trends in sexual orientations in the population over *time and across subpopulations*.

To test this possibility, we expanded the foregoing multinomial logistic model (Table 4). This expanded model added 3-way interaction terms for orientation-by-predictor-by-survey to allow that the two surveys might produce different point estimates of sexual orientation while testing the hypothesis that *trends* over time and across subpopulations in reporting of sexual orientation were statistically equivalent for the two survey programs. This model (Appendix Table A) includes the same five predictor variables shown in Table 4 but adds estimates of 3-way interactions of Survey-by-Predictor-by-Orientation. Estimates of these terms provide tests of the hypotheses that the patterns of Orientation-by-Predictor association found by the two survey programs are statistically equivalent. For example, the *sexual orientation by year by survey* interaction term tests the hypothesis that the two survey programs yield statistically equivalent estimates of *trends* in reported sexual orientation over the years 2014, 2016, and 2018. Similarly, the *sexual orientation by birth cohort by survey* interaction term tests the hypothesis that the two surveys produce statistically equivalent estimates of variation in reported sexual orientations *across birth cohorts*. The remaining 3-way orientation-by-predictor-by-survey terms provide similar tests of statistical significance of the remaining interactions.

The full set of results for this analysis are shown in Appendix Table A, (Since these tests used our expanded model with five additional interaction terms [see Appendix Table A], the estimated relative odds for the main effects are not identical to those reported in Table 4.) It will be seen that none of the 3-way interaction terms were statistically significant. These null results provide an important part of the answer to our basic question about the reproducibility of sexual orientation findings from the two survey programs. We have previously seen that there are statistically significant variations in the two surveys’ point estimates of sexual orientation and that these differences remain even when a variety of sociodemographic controls are applied. Nonetheless, the measurements from these two survey programs do not yield significantly different conclusions about trends in the population’s reported sexual orientations over time or birth cohort nor in the patterns of association between the population’s reported sexual orientations and the other sociodemographic variables included in our model.

## 4. Discussion

Our analysis benefits both from the large, well-executed probability samples used by the GSS and NHIS survey programs and from the well-trained and experienced interview and survey staffs used by these survey programs. While the univariate distributions of social characteristics of the U.S. population are often of minor interest, that has not been the case for estimates of the sexual orientation. Controversies about the size of the U.S. population with non-heterosexual preferences date back, at least, to the 1948 publication of Kinsey et al.’s *Sexual Behavior in the Human Male [23]*. That volume famously estimated that “10 per cent of the males are more or less exclusively homosexual … for at least three years between the ages of 16 and 55. *(p. 651, emphasis in original)*” That figure was both politically controversial and scientifically suspect given the non-probability nature of Kinsey et al.’s sample and other deficiencies in this pioneering work [24].

Subsequent to Kinsey et al.’s companion publication on women [25], relatively little new empirical quantitative research was published on sexual behaviors in the U.S. population. The onset of the HIV/AIDS epidemic in the 1980s began to remedy this deficiency by the end of the 1980s. Reports from the Institute of Medicine and the National Academy of Sciences [26-28] helped bring pressure on the National Institutes of Health to fund the needed research on the sexual behavior of the U.S. population although political opposition slowed these efforts both in the USA [29] and the UK [2].

The subsequent surfeit of survey data in the 1990s and beyond provided much missing information on the variety of sexual behaviors in the U.S. and other populations. The present article advances this effort by providing an evaluation of the reproducibility of an important series of such measurements made by two major survey programs in the USA.

### 4.1 Congruence of Measurements

The data we analyze do not arise from a planned experiment but rather from the fortuitous coincidence of two survey programs measuring the same population characteristic in the first two decades of the 21^st^ century. As we noted previously, the measurement methods used by the two survey programs were not identical in a variety of ways. For example,

- The interview mode varied: video-CASI in the GSS vs. interviewer administration using flashcards in the NHIS;
- The response categories were more extensive in the NHIS including “Something Else”, and an explicit “I don’t know the answer” category;
- The GSS administered a battery of questions on a variety of sexual behaviors, and some respondents opted out of this entire battery of sexual questions. In contrast, missing responses to the NHIS sexual orientation arose only from non-response to this specific question.

Not surprisingly, the point estimates from the two survey programs are not statistically equivalent (Table 2). “Not ascertained,” for example, was a much more common response for the GSS battery of questions (9.5% population estimate) than for the NHIS question (2.2% population estimate). When these responses are purged from the analysis, there still remain some substantial differences. Most notably, the estimated percentage of the population classified as bisexual is more than twice as high in the GSS as in the NHIS (2.21%, se = 0.16% vs. 0.95%, se=0.03%; see Table 2, lower panel). In addition, even when persons refusing the sexual behavior module are excluded from the GSS, the estimated percentage of the population refusing to provide their sexual orientation was almost three times higher in the GSS (1.51%, se = 0.15%) versus the NHIS (0.53%, se=0.02%).

It can be argued that the most important target variables for our analysis is the estimated percentage of the U.S. population that describes itself as gay or bisexual. One common – although perhaps not ideal – approach to this question is to restrict analysis to the three substantive response categories: straight, gay, or bisexual. This analysis strategy (Table 3) does not, however, produce statistically equivalent estimates from the two surveys (p < 0.001), however the estimated percentage of the population classified as gay/lesbian is not significantly different for either the population as a whole (NHIS: 1.59%, se= 0.06% ; and GSS: 1.93%,se=0.17%; p = 0.059) or for males and female considered separately (see Table 3). A relatively large discrepancy arises, however, for the estimated proportion of the U.S. population describing itself as bisexual. Overall, the GSS estimates are more than double those made from the NHIS program (2.90%, se=0.26% vs. 1.03%, se=0.05%). This difference is particularly pronounced for women (GSS: 4.07%, se=0.42% vs. NHIS: 1.42%, se=0.08%).

While the observed *discrepancies* in our survey estimates are less than two percentage points, they are noteworthy because the population characteristics of interest – prevalence of gay and bisexual orientations – are equally small, i.e., 1 to 4 percent. Following a suggestion by England et al. [22], we refined our analysis by adding controls for a variety of factors that are known to influence (reported) sexual orientation. We did find that each of these control variables had a significant statistical association with reporting of sexual orientations. But, the overall discrepancy between the NHIS and GSS survey estimates still remained highly significant (p < 0.001) when these control variables are introduced into our multinomial model. In particular, the GSS population estimates report 1.25 higher relative odds of reporting a gay/lesbian orientation (vs. Straight) than the NHIS and 2.98 higher relative odds of reporting a bisexual (vs. Straight) sexual orientation.

### 4.2 Congruence of Estimated Association between Sexual Orientation and Sociodemographic Predictors

Further analyses indicate that these inter-survey discrepancies do not contaminate conclusions about the associations between sexual orientations and time or our other predictor variables (birth cohort, education, etc.). The formal test of this hypothesis (Appendix Table A) fit a multinomial model that included all five 3-way interactions of sexual orientation-by-predictor-by-survey. None of these interactions were found to be statistically significant.

This result suggests that while the two survey programs *did not yield equivalent point estimates* of the proportion of the USA population reporting different sexual orientations, the survey programs do yield equivalent conclusions about changes in reporting of sexual orientations over time and across subgroups of the USA population defined by birth cohort, gender, education, and race.

## 5. Implications

The foregoing analyses demonstrate that these measurements of sexual orientation in the US population meet a basic test for all scientific measurements. In particular, these two independent series of measurements *do not show* statistically significant divergences in their estimates of the *association* between variations in sexual orientations reported over time or their association with the four other sociodemographic predictors included in our model. The measurements themselves *do*, however, show statistically significant divergences in their point estimates of the proportion of the U.S. population reporting different sexual orientations. In particular, the General Social Survey produces markedly higher estimates of the proportion of the U.S. population labeling themselves as bisexual.

These results have two implications for the use of these data. First, analysts can have some confidence in conclusions regarding both trends over time and the *association* between reported sexual orientation and sociodemographic factors regardless of which of these surveys generated the data. Second, the significant variation observed in point estimates implies that it would be foolhardy to *simply merge* estimates from these two surveys.

### 5.1. Limitations

As noted previously, our analysis benefits both from the large samples surveyed by each survey program and the high-quality field operations that were used. We do note, however, three serious limitations on our conclusions. First, our analysis was restricted to using only five predictor variables: time, gender, birth cohort, education, and race. It is quite possible that we might have reached different conclusions about the congruence of patterns of association if different or additional sociodemographic predictors had been used. Second, our analysis of trends over time is, by necessity, restricted to the three years in which the surveys’ measurements coincided (2014, 2016, 2018). Finally, our analysis does not provide a convincing explanation of why the General Social Survey produced markedly higher estimates of the percent of U.S. adults who report that they are bisexual. The Director of the GSS survey program has suggested that this anomaly may reflect the impact of the GSS survey context, e.g., asking respondents to report on the numbers of male and female sex partners they had since age 18 or the fact that bisexual was the middle category in the GSS question (Tom Smith, personal communication, July 19, 2020). Future research should follow up on these speculations.

## Data Availability

The datasets used in these secondary analyses are publicly available without restriction (https://gss.norc.org/get-the-data and https://nhis.ipums.org/nhis/).

https://nhis.ipums.org/nhis/

## APPENDIX

**Appendix Table A.** Multinomial logistic regression predicting relative risk of reporting a gay or bisexual (vs. straight) orientation as a function of the survey making the measurement, controlling for birth cohort and selected sociodemographic variables plus all survey-by-demographic interactions.

| PREDICTOR | RESPONSE<br>Gay - Lesbian |  |  |  | RESPONSE<br>Bisexual |  |  | *P*<br>*Overall*<br>*Effects (a)* |
| --- | --- | --- | --- | --- | --- | --- | --- | --- |
|  | Coef.<br>Relative Log<br>Odds (a) | Relative<br>Odds<br>Ratio (a) | p |  | Coef.<br>Relative Log<br>Odds (a) | Relative<br>Odds<br>Ratio (a) | p |  |
| **SURVEY** |  |  |  |  |  |  |  |  |
| NHIS | ref |  |  |  | ref |  |  |  |
| GSS | -1.429 | 0.240 | 0.218 |  | 1.398 | 4.048 | 0.061 | 0.039 |
| **GENDER** |  |  |  |  |  |  |  |  |
| Male | ref |  |  |  | ref |  |  |  |
| Female | -0.252 | 0.777 | 0.001 |  | 0.889 | 2.433 | <0.001 | <0.001 |
| **YEAR OF SURVEY** |  |  |  |  |  |  |  |  |
| 2014 | ref |  |  |  | ref |  |  |  |
| 2016 | -0.095 | 0.909 | 0.3 |  | 0.266 | 1.305 | 0.015 |  |
| 2018 | -0.040 | 0.961 | 0.663 |  | 0.447 | 1.563 | <0.001 | 0.002 |
| **BIRTH COHORT** |  |  |  |  |  |  |  |  |
| 1930 & earlier | ref |  |  |  | ref |  |  |  |
| 1940 | 0.394 | 1.483 | 0.073 |  | 0.185 | 1.204 | 0.612 |  |
| 1950 | 0.882 | 2.415 | <0.001 |  | 0.503 | 1.654 | 0.128 |  |
| 1960 | 1.277 | 3.587 | <0.001 |  | 0.780 | 2.182 | 0.012 |  |
| 1970 | 1.138 | 3.119 | <0.001 |  | 1.257 | 3.513 | <0.001 |  |
| 1980 | 1.198 | 3.313 | <0.001 |  | 2.003 | 7.409 | <0.001 |  |
| 1990+* | 1.394 | 4.030 | <0.001 |  | 2.563 | 12.969 | <0.001 | <0.001 |
| **EDUCATION** |  |  |  |  |  |  |  |  |
| < High School Graduate | ref |  |  |  | ref |  |  |  |
| HS Graduate | 0.558 | 1.748 | 0.001 |  | -0.266 | 0.766 | 0.158 |  |
| BA/BS | 0.892 | 2.440 | <0.001 |  | -0.159 | 0.853 | 0.428 |  |
| Post-BA | 1.231 | 3.423 | <0.001 |  | -0.183 | 0.833 | 0.409 | <0.001 |
| **RACE** |  |  |  |  |  |  |  |  |
| White | ref |  |  |  | ref |  |  |  |
| Black | 0.088 | 1.092 | 0.396 |  | -0.349 | 0.705 | 0.012 |  |
| Other | -0.304 | 0.738 | 0.038 |  | -0.329 | 0.720 | 0.126 | 0.126 |
| **INTERACTIONS** |  |  |  |  |  |  |  |  |
| **SURVEY by YEAR** |  |  |  |  |  |  |  |  |
| GSS & 2016 | 0.388 | 1.474 | 0.072 |  | -0.236 | 0.790 | 0.356 |  |
| GSS & 2018 | -0.047 | 0.954 | 0.861 |  | -0.481 | 0.618 | 0.057 | 0.109 |
| **SURVEY by BIRTH COHORT** |  |  |  |  |  |  |  |  |
| GSS & 1940 | 1.879 | 6.544 | 0.074 |  | -0.150 | 0.860 | 0.863 |  |
| GSS & 1950 | 1.311 | 3.708 | 0.229 |  | -0.632 | 0.532 | 0.419 |  |
| GSS & 1960 | 1.060 | 2.888 | 0.317 |  | 0.029 | 1.030 | 0.968 |  |
| GSS & 1970 | 1.327 | 3.772 | 0.211 |  | -0.493 | 0.611 | 0.5 |  |
| GSS & 1980 | 1.784 | 5.951 | 0.087 |  | -0.341 | 0.711 | 0.627 |  |
| GSS & 1990 | 1.476 | 4.374 | 0.17 |  | -0.397 | 0.672 | 0.587 | 0.388 |
| **SURVEY by GENDER** |  |  |  |  |  |  |  |  |
| GSS & Female | 0.102 | 1.108 | 0.628 |  | 0.171 | 1.186 | 0.416 | >0.500 |
| **SURVEY by EDUCATION** |  |  |  |  |  |  |  |  |
| GSS & HS Grad | 0.163 | 1.177 | 0.781 |  | 0.004 | 1.004 | 0.991 |  |
| GSS & BA/BS | -0.059 | 0.942 | 0.919 |  | 0.179 | 1.196 | 0.657 |  |
| GSS & Grad | -0.199 | 0.820 | 0.747 |  | 0.428 | 1.534 | 0.314 | >0.500 |
| **SURVEY by RACE** |  |  |  |  |  |  |  |  |
| GSS & Black | 0.075 | 1.078 | 0.808 |  | 0.383 | 1.467 | 0.138 |  |
| GSS & Other | -0.300 | 0.740 | 0.445 |  | 0.181 | 1.198 | 0.613 | >0.500 |
| **CONS.** | -5.701 | 0.003 | <0.001 |  | -6.616 | 0.001 | <0.001 |  |

NOTE. Coefficients in Appendix Table A vary from those in Table 4 because the multinomial logistic regression model estimated for this table included additional predictor variables (i.e., the sexual *orientation by predictor by survey* interactions) beyond those for the main effects included in the model estimated for Table 4.
(a) Estimates derived by multinomial logistic regression accounting for complex design of samples. Coefficients indicate the estimated relative log odds of reporting Gay-Lesbian (or Bisexual) associated with being in the row category (e.g., female) of the predictor variable compared to the base category (e.g., male) when controlling for other factors in the model. The exponentiated value of this coefficient is the corresponding relative odds ratio. For rare attributes, this value is close to the relative risk ratio.

**Appendix Table B.** Estimates of sexual orientation (a) of U.S. population derived from: (1) complete 2013-18 NHIS adult probability sample, and (2) same sample excluding persons not residing in households.

| Sample |  | Gay/Lesbian | Straight | Bisexual | Unweighted<br>Ns |
| --- | --- | --- | --- | --- | --- |
| **NHIS full**<br>**sample** | est. | **1.64%** | **97.39%** | **0.96%** | **183,020** |
|  | se | **0.04%** | **0.05%** | **0.03%** |  |
| **NHIS**<br>**Household**<br>**Sample (b)** | est. | **1.64%** | **97.41%** | **0.95%** | **181,281** |
|  | se | **0.04%** | **0.05%** | **0.03%** |  |

(a) Analysis excludes persons providing a non-substantive response, e.g., Don't Know.
(b) Excludes NHIS cases whose living quarters were coded as: Not housing unit; Quarters not HU in rooming or boarding house; Unit not permanent in transient hotel, motel, etc.; Unoccupied site for mobile home, trailer or tent; Student quarters in college dormitory; Other group quarter; Other unit - not specified; Other unit - kind unknown; Unknown - Not ascertained. Overall, less than one percent of analysis sample was non-household respondents (unweighted n = 1,739 of 183,020).

